# Epidemiology of chronic pain and opioid use in primary care – a scoping review of big data research

**DOI:** 10.1101/2023.06.12.23291303

**Authors:** Junlin Lin, Hongdian Zhu, Greg Murray, Audrey P Wang

## Abstract

**Objective:** Recent research addressing the opioid use and misuse crisis in patients with chronic non-cancer pain in primary care has focused on traditional cohort studies underpinned by survey data. The advent of electronic health records creates a ‘big data’ opportunity for improving our understanding of the epidemiology of chronic non-cancer pain in primary care and opioid use and misuse. This scoping review aimed to map the chronic non-cancer pain patient population in primary care using big data research, investigating the patient characteristics and opioid prescription patterns.

**Methods:** Searches of primary electronic databases and grey literature, including OVID, CINAHL, and Scopus, were performed from January 1, 2010 to December 2, 2022. The search strategy was restricted to the English language.

**Results:** A total of 1,057 records from databases and 515 records from grey literature were considered. Of these, only three articles met the eligibility criteria, and two articles of these reported an estimated chronic pain prevalence of 3.82% and 10.3% in the primary care setting. Chronic pain patients that presented to primary care providers were predominately female, and common comorbidities were anxiety and depression. An estimated 30% of chronic pain patients used opioids for treatment sourced from general practitioners and family practitioners.

**Conclusion:** The use of big data remains underutilized for investigating the epidemiology of chronic pain and opioid use in primary care. This review calls for a greater focus on pain informatics with big data to improve the accuracy of future clinical chronic pain epidemiology studies.

## Introduction

Chronic non-cancer pain is a growing global health concern, leading to an immense economic burden of $560 billion to $635 billion per year in the United States (U.S.) [1]. Prevalence estimates vary among studies, reportedly from 15% to 50% across European countries and the United States [2–5]. Chronic pain, defined as pain that is present for at least three months [6], is also one of the most common reasons adults seek medical help, and primary care providers are the gateway for chronic pain management [4,7].

Primary care providers frequently prescribe opioid medications as a common chronic pain management strategy. It has been estimated that one in five patients presenting with chronic non-cancer pain in primary care providers receives an opioid prescription [8,9]. This prescribing behavior is typical despite well-documented best-practice guidelines [10–12] and potential iatrogenic harms associated with prescription opioids, including overdose, addiction, and even death [13,14].

Opioid overdose [15,16], misuse [17,18], and abuse [19,20] in the U.S. are endemic and are significant public health challenges. Approximately 4.3 million Americans are engaged in non-medical use of prescription painkillers in any one month (National Survey on Drug Use and Health 2014) [21]. The estimated prevalence of substance abuse disorder is 8.1% in the general population [22], and more than 115 Americans die from opioid overdose daily [21]. In 2008, Benyamin et al. [22] reported that approximately 90% of chronic pain patients received opioids. Furthermore, in 2016, Hwang et al. [23] surveyed and found that 46% of family physicians and general practitioners were unaware that the most common route of abuse was oral prescriptions, and 25% of family physicians who participated in the study were unaware of the potential for opioid diversion to illicit markets [23]. After the Centers for Disease Control and Prevention (CDC) released the 2016 CDC guideline for Prescribing Opioids for Chronic Pain, opioid prescription durations decreased, and non-opioid pain medication prescriptions increased [10,24,25]. However, in 2019, Tong et al. [26] reported that clinicians still found multiple barriers to decreasing the prescriptions of chronic opioids, including contraindications to nonopioid treatment options, restricted availability of adjunctive management approaches, and time constraints [26]. Nevertheless, the latest CDC guideline provided additional guidance on how to assess and mitigate the risks associated with opioid therapy, including patient monitoring and the use of risk stratification tools [27].

Observational studies using large datasets, such as population surveys, electronic health records, and medical claims, can reveal patterns and relationships between variables such as demographic, medical history, lifestyle, psychological, and environmental factors [28–30]. Studies using large datasets can help predict at-risk chronic pain patients [31,32], as the large sample size allows the possibility of extensive subgroup analysis [33,34]; the samples are likely to be more representative of the population [35,36]; linkage opportunities of health data to pharmacy records can offer detailed information for prescription drugs such as type of opioids, dose and supply days [37,38].

Previous reviews of opioids have studied opioid use disorders and opioid misuse in primary care settings [39–41]. For example, younger age groups or female groups in primary care settings are often under-studied with poorly reported prevalence and risk factors [42–45]. To our best knowledge, a review of opioid prescriptions for chronic non-cancer pain patients in primary care providers based on large routinely collected electronic health records, claims, and observational datasets, which we term “big data”, remains lacking.

The aim of this scoping review was to map and describe the characteristics of the chronic non-cancer pain patient population in primary care using big data, investigating the characteristics of those with chronic pain and their opioid prescription patterns. The objectives were to investigate chronic pain patients who received opioid prescriptions for pain management, the settings in which these chronic pain patients were managed, the datasets where the outcomes were reported, and to use the information for future research and strategies.

Our research questions were:

1. What is the prevalence of chronic pain patients in primary care using big datasets?
2. What are the characteristics of chronic pain patients who are present in primary care settings as recorded in big datasets?
3. What are the opioid prescribing patterns for chronic pain patients in primary care? Specifically, we are interested in the following characteristics: supply duration, frequency, and intensity (average morphine milligram equivalents).
4. What are the characteristics of primary care providers who prescribe opioids as the most common chronic pain management strategy for their chronic pain patients?
5. Which source and type of datasets were these chronic pain patients reported in?

## Methodology

This scoping review followed the Preferred Reporting Items for Systematic Reviews and Meta-Analysis extension for scoping reviews (PRISMA-ScR) guidelines [46] and was pre-registered on Open Science Framework at https://osf.io/jtb8w [47]. Searches were conducted in multiple databases to cover interdisciplinary studies, including Medline (Ovid), Embase (Ovid), CINAHL (EBSCO), PsycINFO (Ovid), and Scopus. Grey literature was included. An advanced Google search using a simplified search strategy and targeted websites was employed, including CDC from the U.S., National Health and Medical Research Council (NHMRC) from Australia, GOV.UK from the United Kingdom (U.K.), and Public Health Agency of Canada (PHAC) from Canada. The search was performed on December 2, 2022, restricting the publication date from January 1, 2010 to the performance date. The search strategy, limited to English, used a combination of controlled vocabulary and keywords: 1) chronic non-cancer pain; 2) opioid; 3) primary care provider; 4) big data; 5) cohort studies or cross-sectional studies. Big data in this study was defined as large datasets (sample size ≥ 50,000). Supplementary material listed all search terms used on Medline. We also reviewed reference lists of eligible included studies for additional literature. If the full text was not available for screening, the library access resources were requested. The corresponding authors were also contacted if the required information for this scoping review was not available in both articles and supplementary materials. If authors did not reply within two weeks after the initial enquiry email, a follow-up email was sent. Studies were excluded if authors did not respond within two weeks after the follow-up email. Database search results were exported to EndNote (version 20.2.1) for database grouping. Duplicate removal and screening of titles, abstracts, and full texts of eligible studies were conducted on Covidence, an online systematic review management system.

## Eligibility

Inclusion criteria were as follows: 1) the literature was written in English; 2) participants had chronic non-cancer pain lasting over three months; 3) participants received at least one opioid analgesic for the purpose of chronic pain management; 4) participants presented in primary care providers or outpatient settings; 5) the study sample size ≥ 50,000; 6) observational studies, cohort studies, cross-sectional studies, prospective studies, retrospective studies, and longitudinal studies.

Studies were excluded if: 1) the study described acute pain only or did not provide enough information to determine the presence of chronic pain (i.e., pain lasting for at least three months); 2) the study described postoperative pain which did not develop into chronic pain; 3) the study described chronic pain that was relevant to cancer; 4) the study reported opioid use for purposes other than chronic pain management; 5) the study did not include, or provide enough information to determine the presence of primary care providers; 6) the study described primary care providers who had reasonable reasons to prescribe unlimited opioids for patients (e.g., palliative care); 6) the study population sample size < 50,000; 7) intervention studies, case-control studies, case report studies, and small case series; 8) systematic reviews, scoping reviews, narrative reviews, literature reviews or study protocols.

## Study screening

After removing duplicates, two reviewers (JL and HZ) independently screened titles and abstracts for eligibility using Covidence. The two reviewers assessed full articles for potentially eligible studies. Any disagreements were mediated by a third reviewer (GM) and a fourth reviewer (AW).

Any articles rejected at the full-text screening stage were recorded and categorized by reasons in Figure 1.

**Figure 1.**
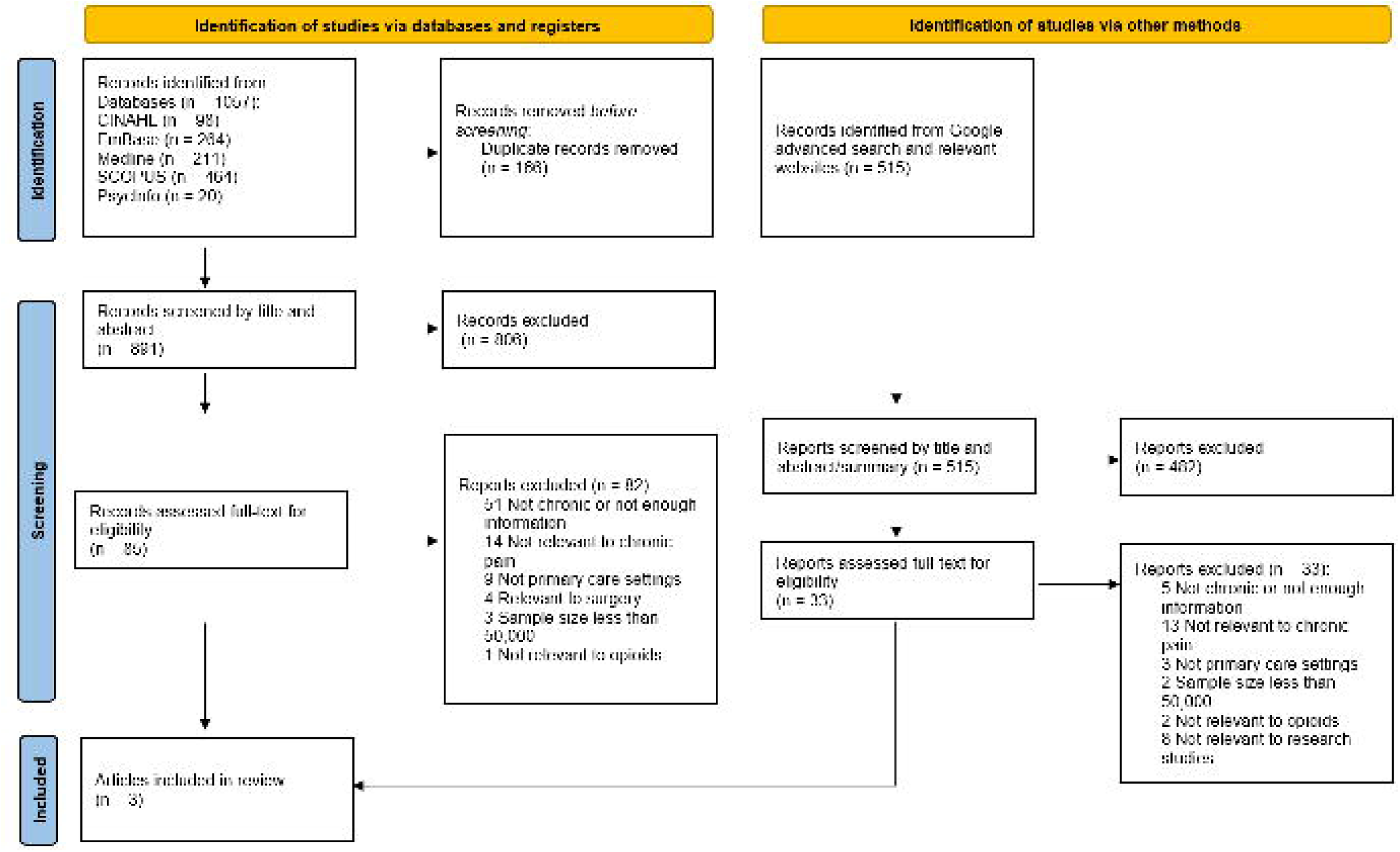
Figure 1: *PRISMA* flowchart

## Data extraction and synthesis

Two reviewers (JL and HZ) independently extracted data using a custom data extraction spreadsheet for each eligible study. Data extraction of eligible studies included descriptive characteristics (authors, year, title, journal, volume, issue, pages, country of the corresponding author, abstract, study design, study setting, and study findings), pain characteristics (definition of pain, pain duration, measures of pain, and pain type/location, prevalence), participant characteristics (sample size, demographics, and number of participants with chronic pain), opioid analgesic characteristics (prevalence, type of opioid analgesic, supply duration, daily morphine milligram equivalent (MME)), primary care provider (e.g., specialty of the provider, number of visits by patients, visit time length), dataset (e.g., source of data, country, type of the dataset). Any disagreements regarding data extraction were mediated by a third reviewer (GM) and a fourth reviewer (AW).

## Quality and risk of bias assessment

The presence of quality and risk of bias assessment utilized the Quality Assessment Tool for Observational Cohort and Cross-sectional Studies published by the National Heart, Lung and Blood Institute [48]. This assessment tool, designed for cohort and cross-sectional studies, evaluated studies from multiple perspectives, including research questions, study population, groups recruited from the same population and uniform eligibility criteria, sample size justification, outcome measures, and others. It returned an overall rating of “Good”, “Fair”, or “Poor” based on appraisals of each part of the study designs. Two individual reviewers assessed the eligible studies independently (JL and HZ), and any disagreements were mediated by a third reviewer (GM) and a fourth reviewer (AW).

## Results

### Study selection

The study selection process is demonstrated in the PRISMA flowchart (Figure 1). The initial results yielded 1057 records across five different databases. First, 166 duplicates were identified and removed both by Covidence and by hand. The remaining 891 records were screened by two reviewers using eligibility criteria, and 85 records were identified for the full-text assessment. Of the 85 articles, seven had conflicts that were resolved by the third reviewer (GM). Eighty-two records, including the seven conflicts, were eventually excluded because they did not meet the eligibility criteria, leaving three eligible studies for the scoping review. Of the excluded records, 51 studies did not have evidence to justify chronic pain, 14 did not study chronic pain, nine were not conducted in primary care settings, four were relevant to surgeries and operations, three had sample sizes <50,000, and one was not relevant to opioids.

Thirty-two of the studies involved in the 85 full-text assessment process applied the International Classification of Diseases, ninth version (ICD-9) codes only, while seven studies applied ICD-10 codes only for the identification of chronic pain. Nineteen studies used both ICD-9 and ICD-10 for chronic pain identifications.

Three peer-reviewed journal articles were included in this scoping review for data extraction. No further articles were identified from hand searching through reference lists. Two authors were contacted, and neither responded.

Among the three eligible studies, both studies by Karmali et al. [49] and Kozma et al. [50] clearly justified and stated chronic pain populations, primary care providers, opioid medications for pain relief, and big data (≥ 50,000). Discussions were required for the inclusion decision of the Lin et al. [51] study; however, agreements were reached by all reviewers that the definition of non-cancer chronic pain can be supported by the one-year follow-up in the study [51].

The grey literature search led to 515 results. Of these, 33 reports underwent full-text assessments, which were mainly from the CDC (U.S.) and GOV.UK (U.K.). No grey literature was eligible for the study.

### Study characteristics and designs

All three eligible studies were conducted in the U.S. and employed medical claim-based datasets for analysis. The study of Karmali et al. reported on chronic musculoskeletal pain [49]; Kozma et al. reported on chronic neck pain, back pain, and osteoarthritis [50], while Lin et al. reported on chronic pain syndrome as well as other pain conditions (back pain and arthritis/joint pain) [51].

Although all three studies were cohort studies, the studies of Karmali et al. [49] and Lin et al. [51] applied a study design using retrospective observational data, whilst the study design by Kozma et al. [50] was a cross-sectional cohort study.

### Background of the included studies

The Karmali et al. [49] study found that increasing the supply of non-pharmacological providers led to a positive impact on the use of non-pharmacological pain treatments and reduced the risk of high-risk opioid prescription patterns. This study focused on individuals aged > 65 years and the potential benefits of increasing access to non-pharmacological pain treatments for older adults with persistent musculoskeletal pain. Their study aimed to “estimate the relationship between the supply of physical therapy and mental health providers and early non-pharmacologic service use with high-risk opioid prescription patterns among Medicare beneficiaries with persistent musculoskeletal pain” in 2007-2014 [49]. High-risk opioid prescription patterns were defined in this study as long-term opioid use (supply duration > 90 days) or high-dose opioid use (daily doses of > 50 morphine milligram equivalents).

The Kozma et al. [50] study investigated if clinical management of nociceptive or neuropathic neck or back pain or osteoarthritis diagnoses were complex and influenced by multiple factors such as patient characteristics, provider involvement, and treatment choices. The study highlighted the need for a comprehensive, patient-centered approach to pain management that considers the patient’s individual needs and preferences. Their study aimed to explore and display descriptive results of various factors that influence pain management, including “patient demographics, comorbidities, office visits, number of different prescribers, overall prescription use in a broad sample of patients with nociceptive or neuropathic neck or back diagnoses or osteoarthritis diagnoses in a commercial insurance population” in 2006 - 2008 [50].

In the Lin et al. [51] study, machine learning algorithms, including logistic regression, least absolute shrinkage and selection operation regression, classification and regression trees, random forests, and gradient boost modeling (GBM), were applied to access the associations of state-wide prescription drug monitoring programs (PDMP) and prescribers’ opioid-related potentially inappropriate prescription (PIP) of opioids for chronic non-cancer pain patients. Their study aimed to “evaluate the U.S. state-wide PDMP effectiveness by examining various PDMP characteristics and their associations with prescribers’ opioid-related PIP practices defined by CDC for chronic non-cancer pain treatment” [51]. The study utilized an extensive all-state medical claims dataset covering 1.8 million beneficiaries from January 2007 to July 2016 and machine learning methods, providing strong statistical power and comprehensive insight into the characteristics of PDMP.

### Chronic pain prevalence and patient characteristics

The study on musculoskeletal pain by Karmali et al. [49] focused exclusively on older individuals who enrolled in the U.S. Medicare Fee-for-Service (FFS) for at least one year and Part D (drug events that contain information about claims for filled prescriptions, including filled date, medication dosage and supply days) for at least six months continuously. The study included patients with two claims diagnosed with musculoskeletal pain > 90 days but < 365 days as the definition of chronic pain. After the selection of the participants, the final cohort was 65,101 patients. No information was provided to allow the calculation of chronic pain prevalence. Of the final cohort, 28.0% were aged 66 to 69 years, which was also the largest age group in the study, and 66.0% of these were female. Of the included 65,101 patients, 84.8% were white; 81.2% of them came from metropolitan counties, and 7.8% of these chronic pain patients also had diagnosed mental health disorders such as anxiety and depression. Of these chronic pain patients, 98.4% presented with arthritis, and 66.3% presented with back pain.

The study on nociceptive or neuropathic neck pain, back pain, or osteoarthritis by Kozma et al. [50] focused on adults aged 18 to 63 years and employed the PharMetrics (Watertown, MA) national managed care database. The study included patients who had at least two pain-relevant claims separated by > 90 days and had at least one oral opioid prescription claim as the chronic pain treatment. Of the population size of 13,163,850 individuals, 85,014 met the eligibility criteria of the present study. Chronic pain prevalence was 10.3% (see Figure 1 in the study, the number with ≥ one study diagnosis between September 1, 2006 and August 31, 2008 observation period, n = 363,152, divided by the number with a valid gender, n = 3,534,443). The average age of the chronic pain patients was 47.8 years old, and 60.4% of these were female. Hypertension was the most common comorbidity among the included chronic pain patients, followed by hyperlipidemia, depression, and diabetes.

The study by Lin et al. [51] investigated PIP for general chronic pain populations of adults > 18 years, using risk factors identified in the 2016 CDC guideline for Prescribing Opioids for Chronic Pain [10]. Chronic pain was defined as multiple general chronic pain, back pain, arthritis, and joint pain relevant to ICD-9 or ICD-10 diagnoses with at least one opioid prescription during a follow-up period > 365 days. The study employed Medicare claims data from Optum De-identified Clinformatics Data Mart and the Prescription Drug Abuse Policy System (PDAPS) database with a population size of 22,264,546 patients. The chronic pain population identified in the study was 851,087 patients, yielding a chronic pain prevalence of 3.82%. The average age of the chronic pain patients was 54.6 years old, and 57.0% of these patients were female. The study reported no further comorbidities.

### Opioid medications

In the Karmali et al. study [49], 13.2% filled an opioid prescription during the first three months of the persistent pain episode, and 31.3% of the patients filled at least one opioid prescription during the follow-up period. The average opioid supply for the study population was 28.2 days, and the average daily dose of opioids was 43.6 MME per patient.

In the Kozma et al. study [50], the average number of pain-related prescriptions for each chronic pain patient was 8.8. Over 80% of patients filled weak opioids (WHO pain relief ladder rung two for moderate pain, e.g., tramadol, codeine, dihydrocodeine), and over 30% of patients filled strong opioids (WHO pain relief ladder rung three for severe pain, e.g., morphine, fentanyl, buprenorphine, oxycodone, hydromorphone). No information was provided about the duration of opioid supply and the average MME.

In the Lin et al. study [51], there were 829,723 out of 851,087 chronic pain patients who filled at least one prescription, leading to a high opioid prescription rate of 97.49%. It was estimated that 70.3% of claims involved prescriptions for an opioid supply over seven days. Further, 22.8% of all opioid prescription claims for chronic pain patients were > 50 MME/day, while 8.9% of all opioid prescription claims for chronic pain patients were > 90 MME/day. Moreover, 10.3% of opioids were co-prescribed with benzodiazepines within seven days of the initial prescription date of opioids. These two descriptors, the seven-day opioid supply and the co-prescribing of benzodiazepines, were defined in the 2016 CDC Opioid Prescribing Guidelines as high-risk prescribing patterns [10].

### The characteristics of primary care providers

In the Karmali et al. study [49], the primary care providers included the county-level annual supply of general practice, geriatric specialists, surgeons, physical medicine and rehabilitation specialists, nurse practitioners, physician assistants, and pharmacists. The primary care providers’ usage rate was 6.8 per 10,000 people per county.

In the Kozma et al. study [50], the primary care providers included general and family practice and internal medicine physicians. The average number of office visits per patient during the observation period was 32.0. The most frequented providers were general and family practice (39.8% of all visits of chronic pain patients), followed by internal medicine physicians (16.9%).

The primary care provider specialty, average office visits, and average MME prescribed by providers were not reported in the Lin et al. study [51]. However, the study documented that 60.9% of these opioid prescriptions were from primary care providers.

### Big data analysis methods

The Karmali et al. study [49] employed a logistic regression model to estimate the odds of high-risk opioid prescription patterns for each standard deviation increase in the supply of non-pharmacological providers. In this study, non-pharmacological providers were defined as physical therapy and mental health providers. The model contained variables such as non-pharmacological pain treatments, demographic characteristics, and comorbidities. The results showed that areas with a higher supply of non-pharmacological providers were associated with a lower prevalence of high-risk opioid prescription patterns.

The Kozma et al. [50] study employed no logistic regression model or machine learning algorithms.

The Lin et al. study [51] applied machine learning algorithms for the following tasks: “1) to determine which machine learning model has the best performance of opioid-related PIPs predictions; 2) to identify the most important PDMP characteristics by both regression-based models (i.e., the characteristics associated with opioid related PIPs) and tree-based models (i.e., the importance of characteristics); and 3) to determine the magnitudes and directions of the important PDMP characteristics on opioid-related PIPs.” GBM outperformed other classifications with an accuracy of over 0.71 for all types of high-risk prescribing patterns.

### Quality assessment

All three studies underwent quality appraisal using the Quality Assessment Tool for Observational Cohort and Cross-sectional Studies published by the National Heart, Lung and Blood Institute of the NIH (National Institutes of Health). Both reviewers answered assessment questions for these three studies and reached an agreement on the overall rating, as shown in Table 1.

**Table 1:**
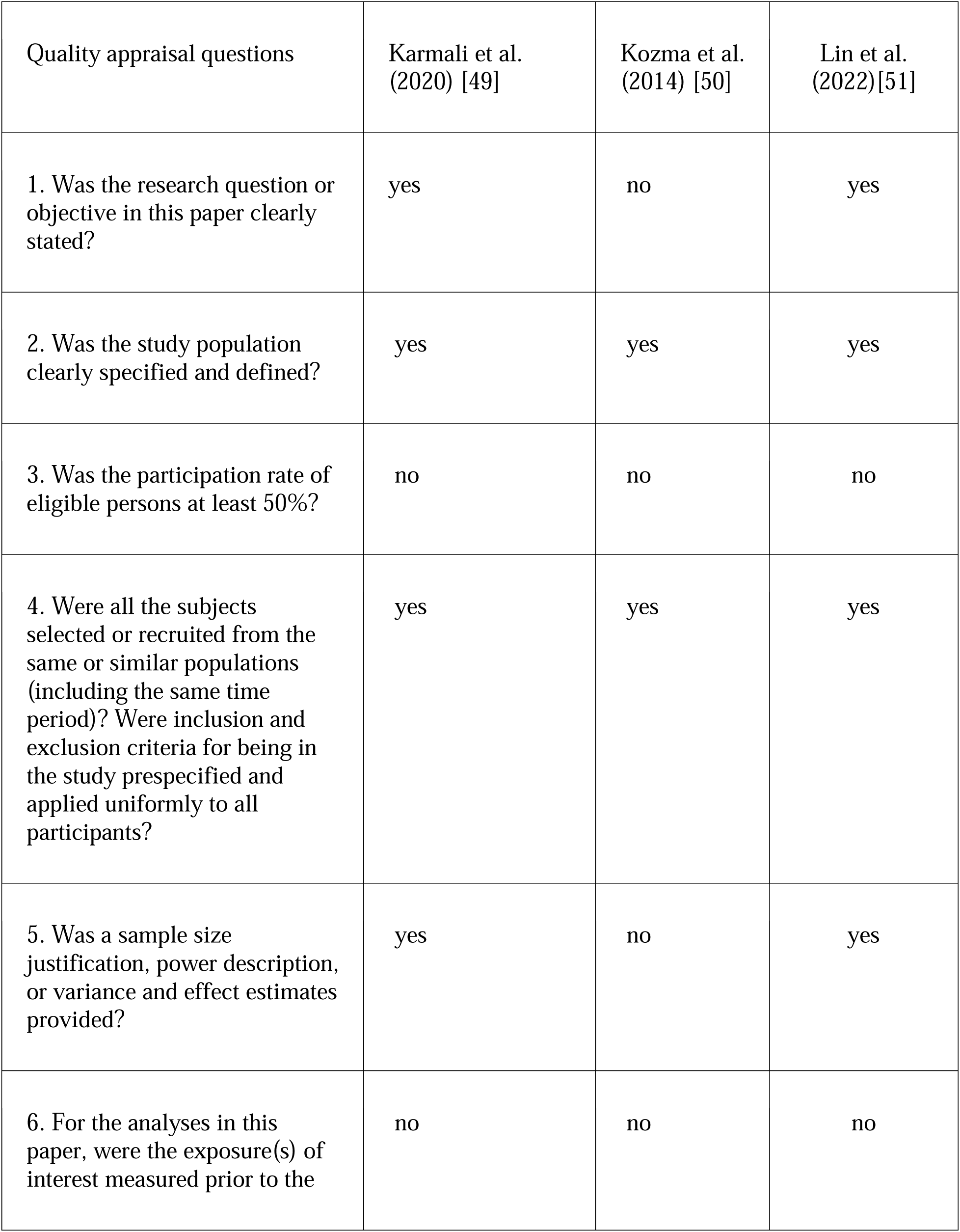
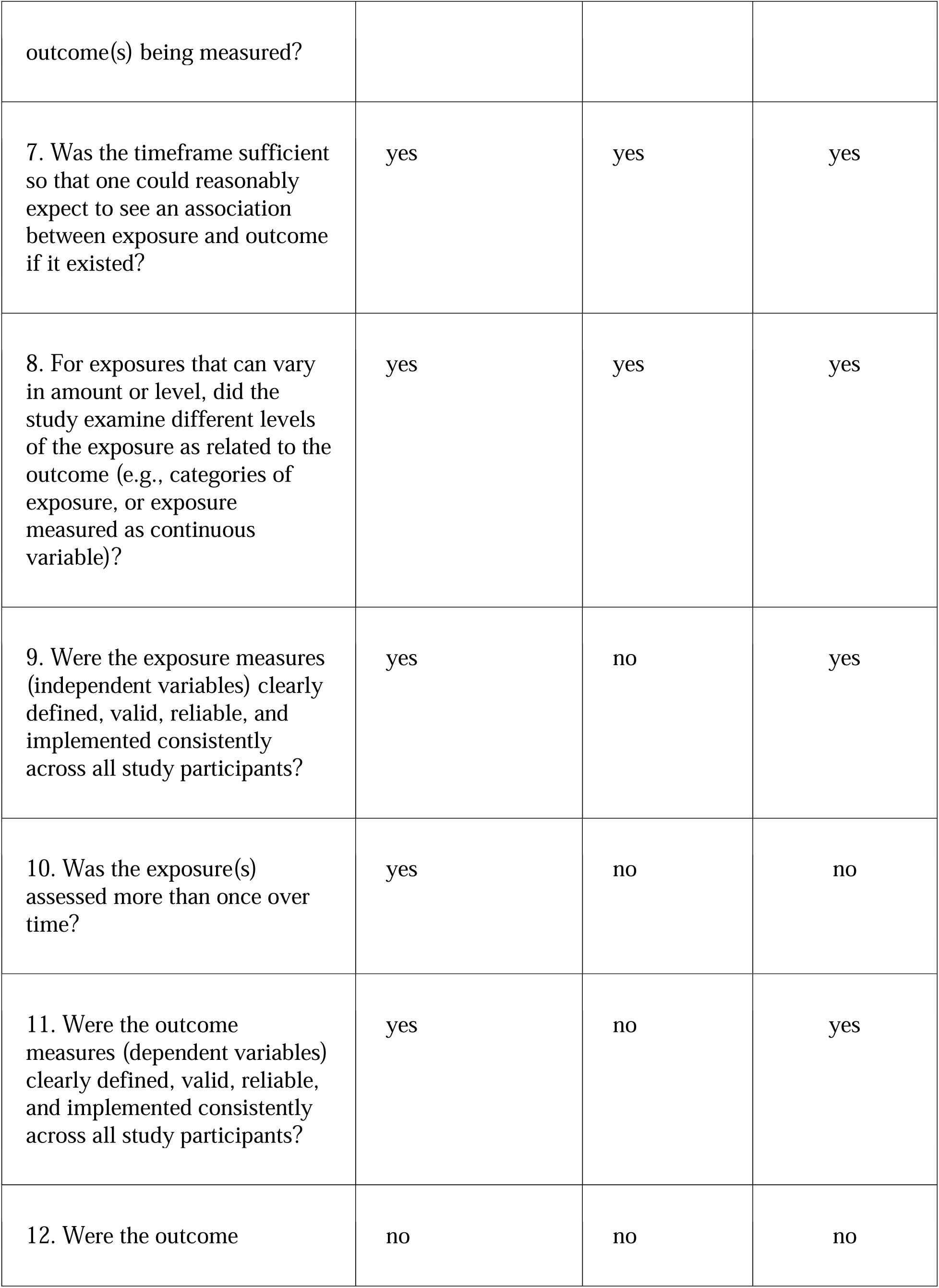
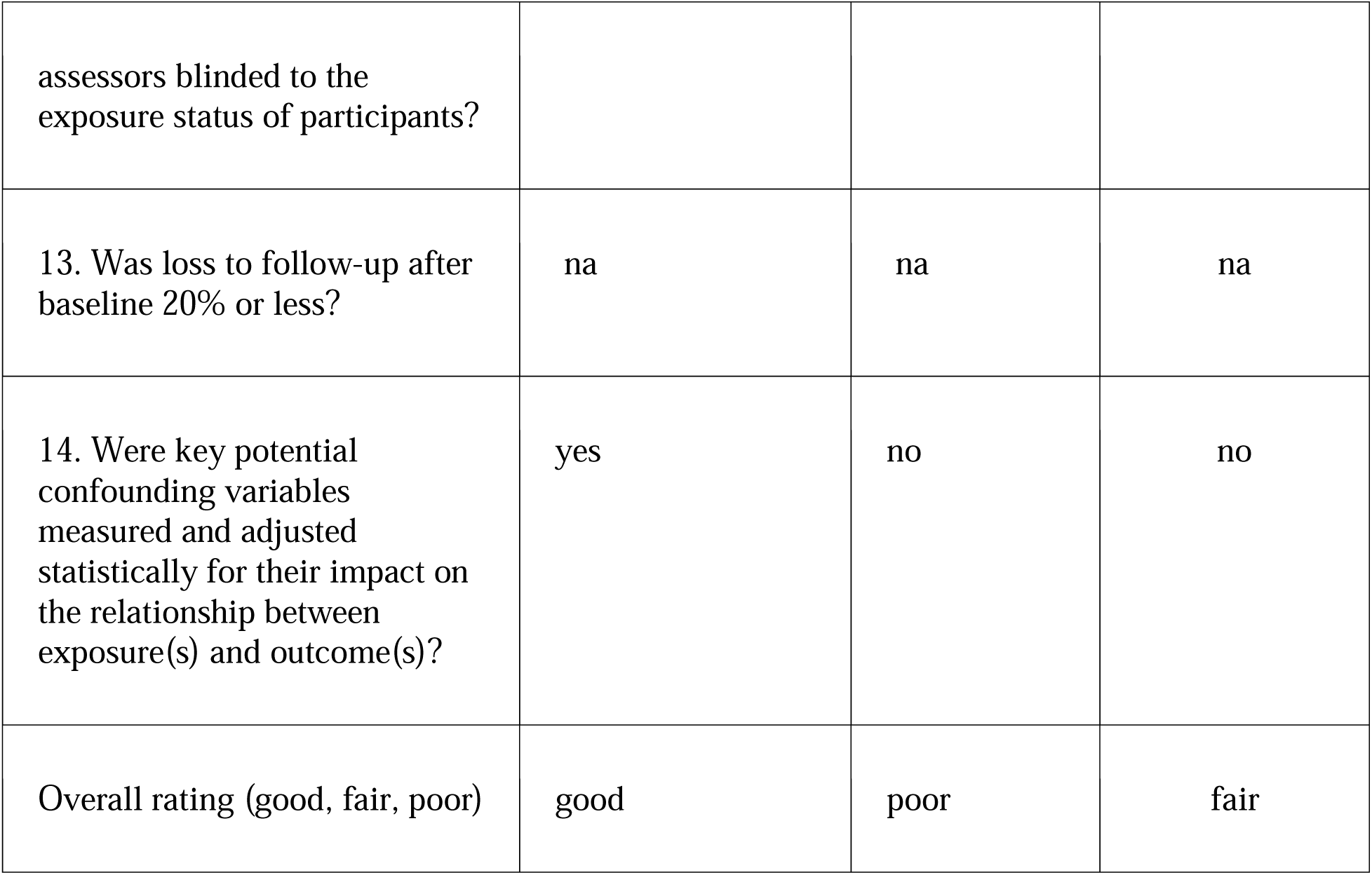
quality assessment of the studies

## Discussion

There is a demonstrable research gap between previous chronic pain research and this new area, which we describe as pain informatics using big data in progressing our understanding of chronic pain epidemiology. Among the original 891 records imported for screening, only three peer-reviewed articles were eligible for the scoping review using the criteria. Although all three studies included the perspectives of chronic pain, opioid use, primary care, and the employment of big data analysis, none was able to comprehensively address all of our research questions.

The Karmali et al. study [49] employed a large dataset of Medicare beneficiaries, which supports the study of a representative population. However, retrospective studies are at a disadvantage as they are unable to establish causality. The comprehensive descriptive analysis performed in the Kozma et al. study [50] provided summaries of chronic pain management across four cohorts. However, the limitation of cross-sectional studies was that the temporal link between the outcome and the exposure could not be determined because both are examined at the same time, which can lead to the failure to draw valid conclusions as to the association between a risk factor and health outcomes. The Lin et al. study [51] utilized an extensive all-state medical claims dataset covering 1.8 million beneficiaries and applied multiple machine-learning methods, providing a much larger sample size and allowing a comprehensive data linkage insight into the characteristics of PDMP. However, the Lin et al. study’s [51] primary unit was claims per patient rather than patient, which may potentially lead to less relevant information regarding the overall chronic pain population.

Our review study was not able to estimate the chronic pain prevalence in primary care using big data research. All three studies did not report the chronic pain prevalence directly in the study but were estimated from two of the studies as 10.3% (nociceptive or neuropathic neck pain, back pain, or osteoarthritis in adult U.S. individuals 18 to 63 years) [50] and 3.82% (chronic pain) [51]. Both estimates were significantly below the 20.4% chronic pain prevalence reported by the CDC in 2018 [3]. One potential explanation for the difference in the estimated prevalence of chronic pain between the studies and the CDC’s report is the variation in the study inclusion criteria. For example, Kozma and Lin’s studies have included only specific types of chronic pain diagnoses, such as osteoarthritis and low back pain [50,51], while the CDC’s report provided a general summary of any kind of chronic pain with a time-based limitation, such as “in the last three months” [3]. The use of diagnostic codes in these two studies could also have impacted the prevalence estimates, as it may have excluded individuals with chronic pain who did not have a formal diagnosis. These variations suggest that the prevalence of chronic pain in the U.S. is still a matter of ongoing research and may be affected by various factors.

Our review study found that chronic pain patients that presented at primary care providers were disproportionately female in all three studies. This is representative of current chronic pain research, as females constituted 57.6% on average of the chronic pain population from the three studies [26,52–54]. One of the most common comorbidities mentioned by Karmali et al. [49] and Kozma et al. studies [50] was mental health disorders such as anxiety and depression. This, again, is reflective of the current chronic pain research [26,55–57].

Our review study found that an estimated 30% of chronic pain patients used opioids for treatment. Karmali et al. [49] reported that the average number of days of opioid supply was 28.2 days, and the average MME was 43.6 mg/day. They indicated that opioid prescription filling was 31.3% during the one-year follow-up period. The Kozma et al. study [50] reported that > 30% of patients had pain relief prescriptions that were considered strong opioids as classified by the WHO pain ladder. Notably, Lin et al. [51] demonstrated an estimated 23% of claims with MME > 50 mg/day and 9% of claims with MME > 90 mg/day. Considering the overlapping study period of these studies (2008-2013), the fact is that the opioid prescribing patterns of primary care providers were a concerning public health problem before the release of the 2016 CDC opioid prescribing guideline. Benyamin et al. [22] reported that 90% of chronic pain patients received opioids in 2008, whilst a later study in 2014 using National Ambulatory Medical Care Survey data in the U.S. reported that 36.4% of chronic pain patients received opioids [58]. Our review study, covering 2010 to 2022 big data research, found that an estimated 30% of chronic pain patients received opioids for treatment sourced from general practitioners and family practitioners. There appears to be some progress since the CDC released its guidelines in 2016 for reducing the usage of opioids. Still, ultimately an even more significant reduction in receiving opioids by chronic pain patients should be balanced with an increase in the uptake of non-pharmacological treatments.

Our review study was not able to report the characteristics of primary care providers who prescribed opioids as the most common chronic pain management strategy for their chronic pain patients, as none of the three studies provided much information on it. However, both the Karmali et al. [49] and the Kozma et al. [50] studies reported that the most frequently visited primary care providers were general practitioners and family practitioners. In the Karmali et al. study [49], the average primary care provider supply was 6.8 per 10,000 people per county. Further, it was reported that primary care providers like general and family practice constituted 39.8% of the total office visits by chronic pain patients in the study conducted by Kozma et al. [50]. Additionally, the Lin et al. study [51] showed that primary care providers prescribed 60.9% of opioid prescriptions, more than any other specialties in the U.S. healthcare system.

Our review study found that all three studies used medical claims datasets. All three databases were patient-centric and de-identified, associated with corresponding pharmacy and provider claims information.

Although these three eligible studies only partially addressed our proposed research questions, valuable insights into the current management of chronic pain in primary care settings from different perspectives have highlighted the complexity of the issue and the need for a comprehensive, patient-centered approach to pain management in primary care settings.

Our study is the first known study reviewing the utilization of big data in chronic pain and opioid use in the primary care setting. This review adds to the research evidence base, providing an estimate of the population chronic pain prevalence and clinical opioid prescriptions for pain management in primary care settings using big data. Through a comprehensive review, we found only three eligible studies, revealing a substantial lack of high-quality research in this domain. Leveraging the power of big data in pain informatics is promising, as it mitigates the risk of biases often associated with traditional questionnaires and surveys, including selection bias [59,60], recall bias [61,62], and non-response bias [63–65]. By harnessing big data, studies in chronic pain research can attain greater representativeness, yielding findings that can be more applicable to the broader population and potentially foster more evidence-based clinical practices. Moreover, our study investigated opioid prescribing patterns for chronic pain management in the primary care setting. This is a potentially vital target for future policies to improve patient care regarding chronic pain and the opioid epidemic.

Our review study also has several limitations. Specifically, we limited our search to studies after 2010 and English language only. Furthermore, the inclusion and exclusion criteria of chronic pain were strictly defined as pain symptoms lasting > 90 days according to IASP [6], while most studies failed to demonstrate this IASP eligibility criterion in their study and often used less stringent definitions.

The varied reporting of chronic pain prevalence indicates that future research should be carried out using big datasets. To improve our understanding of the global health concern of chronic pain, the prevalence of chronic pain is required to provide public health surveillance of the scale of the problem. Compared to estimating the population chronic pain prevalence via a small sample size of < 50,000 patients, large real-world datasets can draw from a larger and more diverse population, thus providing a more comprehensive and representative sample of the general chronic pain population. A disadvantage of medical claims data is the inconsistency of chronic pain diagnostic coding systems. Because ICD-9 and ICD-10 codes lacked a systematic categorization of chronic pain, it was difficult to confirm that patients suffered from chronic pain based on one or multiple specific diagnostic codes [66]. Future research should aim to maintain the consistency of the chronic pain diagnostic coding system, which has been identified in the IASP ICD-11 seminar paper [67]. The lack of investigation into prescription opioid frequency is because studies do not mainly take patients as the primary unit in clinical epidemiology. Therefore, counting the basic unit as patient versus claim can be the main challenge of studying opioid prescription frequency.

## Conclusion

Much remains unknown regarding chronic pain population prevalence, opioid prescribing patterns, and primary care provider characteristics using big data and requires further research. However, the topic of pain informatics is an increasingly promising area of research as research insights have comparatively increased from a decade ago, primarily due to the digitization of patient records. This review showed a chronic pain prevalence of 3.82% and 10.3% in primary care settings, according to two eligible articles, lower than the current estimates of 15%-50%. This review also found that chronic pain patients that presented at primary care providers were predominately female, and the most common comorbidities present were anxiety and depression. A larger focus on understanding chronic pain identification and prediction using big data during this developmental stage of pain informatics may potentially lead to improved treatment options and better long-term outcomes.

## Data Availability

All data produced are available online at https://osf.io/jtb8w

## Acknowledgment

We would like to express our sincere gratitude to Tess Aitken and Kanchana Ekanayake, Academic Liaison Librarian of The University of Sydney library, for their assistance in our scoping review. Their guidance has been instrumental in enhancing the quality and efficiency of our research.

## Supplementary material

A comprehensive list of the search terms used in the Medline database to retrieve relevant articles for the study was included as the supplementary material.

