## supplementary material for "Epidemiology of chronic pain and opioid use in primary care – a scoping review of big data research"

**Database: Ovid MEDLINE(R) ALL <1946 to December 2, 2022>**
**Search Strategy:**
**1**  (chronic adj3 pain*).mp. (86405)
**2**  ((noncancer* or non-cancer* or chronic* or recurrent or persist* or non-malign*) adj3 pain).mp. (102783)
**3**  Chronic Pain/ (22075)
**4**  Osteoarthritis/ or osteoarthrit*.mp. or osteo-arthritis.mp. (111868)
**5**  degenerative arthrit*.mp. (1429)
**6**  Arthritis, Rheumatoid/ (109427)
**7**  Neuralgia/ or neuralg*.mp. (33226)
**8**  Diabetic Neuropathies/ or (neuropath* adj3 (pain* or diabet*)).mp. (49506)
**9**  zoster.mp. (23974)
**10**  Irritable Bowel Syndrome/ or (IBS or irritable colon or irritable bowel).mp. (20216)
**11**  Fibromyalgia/ or fibromyalg*.mp. (13925)
**12**  complex regional pain syndromes/ or causalgia/ or reflex sympathetic dystrophy/ or (complex regional pain syndromes or causalgia).mp. (6228)
**13**  Pain, Intractable/ (6370)
**14**  Hyperalgesia/ (13874)
**15**  back pain/ or low back pain/ (44344)
**16**  Radiculopathy/ or radiculopathy.mp. (10747)
**17**  musculoskeletal pain/ (4516)
**18**  Arthralgia/ (9816)
**19**  Migraine Disorders/ or migraine.mp. (45184)
**20**  Headache Disorders/ or headache/ or headache*.mp. (111362)
**21**  Cervicalgia.mp. or Neck Pain/ (8586)
**22**  Temporomandibular Joint Dysfunction Syndrome/ or TMJ* pain*.mp. (5564)
**23**  whiplash.mp. or whiplash injury/ (4213)
**24**  Cumulative Trauma Disorders/ (4620)
**25**  (trigonitis or trigonites).mp. (48)
**26**  Interstitial cystitis.mp. or Cystitis, Interstitial/ (4232)
**27**  Vulvar vestibulitis.mp. or Vulvar Vestibulitis/ (326)
**28**  vulvodynia.mp. or Vulvodynia/ (1044)
**29**  dorsalgia.mp. (126)
**30**  Endometriosis/ or Endometriosis.mp. (32876)
**31**  (pelvic pain or perineal pain).mp. (14339)
**32**  dysmenorrhea.mp. or Dysmenorrhea/ (7623)
**33**  dyspareunia.mp. or Dyspareunia/ (5750)
**34**  (backache* or polyarthralgi* or arthrodyni* or myalgi* or myodyni* or ischialgi* or crps or rachialgi*).ab,ti. (19466)
**35**  ((back or discogen* or bone or musculoskelet* or muscle* or skelet* or spinal or spine or vertebra* or joint* or arthritis or Intestin* or neuropath* or neck or cervical* or head or facial* or complex or radicular or cervicobrachi* or orofacial or somatic or shoulder* or knee* or hip or hips) adj3 pain).mp. (206608)
**36**  1 or 2 or 3 or 4 or 5 or 6 or 7 or 8 or 9 or 10 or 11 or 12 or 13 or 14 or 15 or 16 or 17 or 18 or 19 or 20 or 21 or 22 or 23 or 24 or 25 or 26 or 27 or 28 or 29 or 30 or 31 or 32 or 33 or 34 or 35 (759072)
**37**  Analgesics, Opioid/ or (opioid* or opiate*).mp. (159409)
**38**  Narcotics/ or narcotic*.mp. (65674)
**39**  (alfentanil or alphaprodine or beta-casomorphin$ or buprenorphine or carfentanil or codeine or deltorphin or dextromethorphan or dezocine or dihydrocodeine or dihydromorphine or enkephalin$ or ethylketocyclazocine or ethylmorphine or etorphine or fentanyl or heroin or hydrocodone or hydromorphone or ketobemidone or levorphanol or lofentanil or meperidine or meptazinol or methadone or methadyl acetate or morphine or nalbuphine or opium or oxycodone or oxymorphone or pentazocine or phenazocine or phenoperidine or pirinitramide or promedol or propoxyphene or remifentanil or sufentanil or tilidine or tapentadol).mp. (170566)
**40**  (adolonta or Anpec or Ardinex or Asimadoline or Alvimopam or amadol or biodalgic or biokanol or Codinovo or contramal or Demerol or Dicodid or Dihydrocodeinone or dihydromorphinone or dihydrohydroxycodeinone or dihydrone or dilaudid or dinarkon or dolsin or dolosal or dolin or dolantin or dolargan or dolcontral or duramorph or duromorph or duragesic or durogesic or eucodal or Fedotzine or Fentanest or Fentora or Fortral or Hycodan or Hycon or Hydrocodone or Hydrocodeinonebitartrate or hydromorphon or hydroxycodeinon or isocodeine or isonipecain or jutadol or laudacon or l dromoran or levodroman or levorphan or levo-dromoran or levodromoran or lexir or lidol or lydol or morfin or morfine or morphia or morphin or morphinium or morphinene or morphium or ms contin or n-methylmorphine or n methylmorphine or nobligan or numorphan or oramorph or oxycodeinon or oxiconum or oxycone or oxycontin or palladone or pancodine or pethidine or phentanyl or prontofort or robidone or skenan or sublimaze or sulfentanyl or sulfentanil or sufenta or takadol or talwin or theocodin or tramadol or tramadolhameln or tramadolor or tramadura or tramagetic or tramagit or tramake or tramal or tramex or tramundin or trasedal or theradol or tiral or topalgic or tradol or tradolpuren or tradonal or tralgiol or tramadorsch or tramadin or tramadoc or ultram or zamudol or zumalgic or zydol or zytram).mp. (12474)
**41**  37 or 38 or 39 or 40 (285116)
**42**  Primary Health Care/ or (primary adj3 provider*).mp. (101220)
**43**  (practitioner* or physician* or clinician*).mp. (1063497)
**44**  General Practice/ or general practi*.mp. or General Practitioners/ (102391)
**45**  Physicians, Family/ or Family Practice/ or family practi*.mp. (85208)
**46**  Nurse Practitioners/ or nurse practitioner*.mp. (25265)
**47**  Internal Medicine.mp. or Internal Medicine/ (38510)
**48**  (gynecologist* or obstetrician* or (obgyn or ob-gyn)).mp. (24774)
**49**  Chiropractic/ or chiropractor*.mp. (4641)
**50**  Physical Therapists/ or Physical Therapy Specialty/ or physical therap*.mp. (64678)
**51**  (an?esthesiology adj3 pain medicine).mp. or Pain Management/ (40679)
**52**  42 or 43 or 44 or 45 or 46 or 47 or 48 or 49 or 50 or 51 (1314969)
**53**  Diagnosis, Computer-Assisted/ or Artificial Intelligence/ or Machine Learning/ or Algorithms/ or Pattern Recognition, Automated/ (367402)
**54**  Data Mining/ or Big Data/ or Information Management/ or data analysis.mp. or Data Analysis/ (119321)
**55**  ((medic* adj3 claim*) or (Medicaid or Medicare)).mp. (107751)
**56**  medical record*.mp. or Medical Records/ or Electronic Health Records/ or electronic health record*.mp. or Medical Records Systems, Computerized/ or EHR*.mp. (278657)
**57**  53 or 54 or 55 or 56 (846370)
**58**  Retrospective Studies/ or Prospective Studies/ or observational stud*.mp. (1883815)
**59**  Cross-Sectional Studies/ or cross sectional stud*.mp. (541729)
**60**  Longitudinal Studies/ or longitudinal stud*.mp. (209122)
**61**  Cohort Studies/ or cohort stud*.mp. (527922)
**62**  58 or 59 or 60 or 61 (2739154)
**63**  36 and 41 and 52 and 57 (606)
**64**  62 and 63 (304)
**65**  limit 64 to (english language and yr="2010 - 2023") (277)
**66**  Randomized Controlled Trials as Topic/ or rct*.mp. or intervention stud*.mp. or Clinical Trials as Topic/ or clinical trial*.mp. or control* stud*.mp. or random* control* trial*.mp. (2057188)
**67**  (case stud* or case serie* or case report*).mp. or Case Reports/ (2617911)
**68**  "Systematic Review"/ or "Review Literature as Topic"/ or systematic review*.mp. or scoping review*.mp. or literature review*.mp. or narrative review*.mp. (475412)
**69**  Postoperative Complications/ or (post adj3 operati*).mp. or (post adj3 surg*).mp. or (after adj3 operati*).mp. or (after adj3 surg*).mp. or postsurg*.mp. or postoperati*.mp. (1297968)
**70**  66 or 67 or 68 (4929557)
**71**  69 or 70 (5821924)
**72**  65 not 71 (211)
